# Reduced egocentricity after psilocybin in patients with alcohol use disorder: a pilot randomized controlled trial

**DOI:** 10.64898/2026.09.26.26364086

**Authors:** Henryk Bukowski, Philippe de Timary, Dahbia Belahda, Noé Igounenc, Thibault Mura, Chris Serrand, Amandine Luquiens

**Author notes:** Corresponding author: Amandine Luquiens, Univ. Montpellier, Department of Addiction, CHU Nîmes Nîmes, France.

## Abstract

**Background:** Social difficulties in alcohol use disorder (AUD) have been linked to alterations in social cognition, including increased egocentric bias. Psychedelics such as psilocybin have been proposed to modulate self-related processing, but their effects on specific socio-cognitive mechanisms remain unclear.

**Methods:** In this randomized controlled pilot study, recently detoxified individuals with AUD with persistent depressive symptoms completed a visual perspective-taking task before and after a psilocybin session (25 mg dose vs active placebo 1 mg psilocybin).

**Results:** The 25 mg psilocybin group showed a significant reduction in egocentric bias and egocentric superiority on reaction times, with no effect on altercentric bias nor on accuracy. Between-group comparisons confirmed greater reductions in the high-dose relative to the low-dose group. The high-dose group reported more intense subjective psychedelic experiences, and greater subjective intensity was associated with lower egocentric superiority in accuracy.

**Conclusions:** These findings provide preliminary evidence that psilocybin may selectively reduce egocentric interference in individuals with AUD, suggesting a decrease in egocentricity rather than enhanced sensitivity to others. Although replication in larger samples is needed, reduced egocentricity may represent one mechanism through which psilocybin influences social cognition in AUD.

**Clinical trial registration:** ClinicalTrials.gov, https://clinicaltrials.gov/study/NCT06235411, NCT06235411.

## 1. Introduction

Substance use and addictive disorders constitute a major public health burden, with alcohol use alone representing one of the leading risk factors for mortality worldwide among individuals aged 15–49 years and a substantial contributor to disability-adjusted life-years (Griswold et al., 2018; World Health Organization, 2019). Although detoxification and standard psychosocial interventions can effectively reduce acute symptoms such as craving, anxiety and depressive affect (de Timary et al., 2013; Petit et al., 2017), long-term outcomes remain highly variable, with relapse rates remaining substantial. Importantly, sustained abstinence critically depends on the quality of everyday social interactions and support networks (Kaskutas et al., 2002; Kelly et al., 2009, 2012). These interactions rely on intact social-cognitive processes, suggesting that impairments in how individuals represent and respond to others may play a key role in addiction trajectories.

A growing body of evidence indicates that individuals with severe alcohol use disorder exhibit impairments in social cognition, defined as the processes supporting the perception and interpretation of others’ behaviours, thoughts and emotions (Bora & Zorlu, 2017; Pabst et al., 2022). These impairments have often been described as reflecting deficits in theory of mind, that is, difficulties in considering other people’s perspectives and accurately inferring their mental states (Bosco et al., 2014; Uekermann et al., 2007). However, tasks used to assess theory of mind typically involve multiple cognitive processes, making it difficult to identify the specific mechanisms underlying these deficits. A growing alternative view is that many of these findings may be more parsimoniously explained by an increased egocentric bias, reflecting the combined influence of heightened self-centred processing and reduced ability to inhibit one’s own perspective when it conflicts with that of others (Epley et al., 2004; Leslie et al., 2004; Samson & Apperly, 2010). In this framework, apparent deficits in mental state understanding do not necessarily reflect a primary impairment in inferential abilities, but rather a difficulty in overcoming the dominance of self-related representations. This interpretation is consistent with broader evidence in alcohol use disorder pointing to both increased self-focus and impairments in inhibitory control, two processes that jointly contribute to egocentric interference in social cognition (Maurage et al., 2015; Pabst et al., 2021).

The visual perspective-taking (VPT) task developed by Samson and colleagues (2010) provides a direct and process-specific way to quantify the egocentric bias. In this paradigm, participants are required to judge what another agent can see while ignoring their own potentially conflicting perspective (Samson et al., 2010). Performance costs when judging the other’s perspective in the presence of conflicting self-related information provide a direct index of egocentric interference, whereas interference from the other’s perspective when judging one’s own perspective reflects altercentric bias. Further, these two biases can be subtracted from one to another to index the egocentric superiority and cancel out their interference resolution shared component. Hence, this paradigm allows the decomposition of social-cognitive performance into distinct and quantifiable indexes, offering a more process-specific approach tool to investigate the mechanisms underlying socio-cognitive impairments in alcohol use disorder than broader theory-of-mind tasks (Bukowski & Samson, 2017).

Recent years have also seen renewed interest in the therapeutic potential of psychedelics, including psilocybin, for the treatment of alcohol use disorder (Bogenschutz et al., 2022; Rieser et al., 2025). Clinical studies suggest that psilocybin-assisted interventions can produce sustained reductions in alcohol consumption and related symptoms, yet the cognitive mechanisms underlying these effects remain poorly understood (Luquiens et al., 2025). A prominent hypothesis is that psychedelics act by altering self-related processing, including reducing rigid self-focus and increasing cognitive flexibility (Aday, Carhart-Harris, & Woolley, 2023). From this perspective, psilocybin may reduce the dominance of self-related representations in situations of conflict, thereby facilitating a more balanced integration of internal and external information.

Extending this hypothesis to social cognition, psilocybin may specifically reduce egocentric bias by decreasing the interference of one’s own perspective when processing others’ viewpoints. Such an effect would be particularly relevant in alcohol use disorder, where heightened self-focused attention leading to negative thinking and impaired inhibition are central features (Mansueto et al., 2024; Maurage et al., 2015). By improving the ability to overcome self-related interference, psilocybin could enhance social-cognitive functioning and, in turn, contribute to better interpersonal functioning and treatment outcomes.

The present study tested this hypothesis using a VPT task administered to patients with alcohol use disorder (AUD) participating in a randomized controlled trial comparing a high dose (25 mg) versus a low dose (1 mg) of psilocybin. Participants completed the task at baseline and after the first dosing session. We hypothesized that psilocybin would reduce egocentric bias, and that this reduction would be greater in the high-dose group compared to the low-dose group.

## 2. Material and methods

### 2.1. Transparency and ethics

Due to GDPR regulations and the sensitive nature of clinical data, participant data are not publicly available but may be made available from the corresponding author upon reasonable request and subject to ethical approval and applicable data protection regulations. This study was conducted in accordance with the principles of the Declaration of Helsinki and adhered to Good Clinical Practice guidelines. It was approved by the French Ethics Committee (Comité de Protection des Personnes) and the French National Agency for the Safety of Medicines and Health Products (ANSM) (n°2023-506647-40-02) and preregistered on clinicaltrials.gov (NCT06235411). All participants provided written informed consent.

### 2.2. Sample

This study is a cognitive analysis of the Psilocybin Alcohol Depression (PAD) trial, a prospective, single-centre, double-blind, randomized controlled pilot study investigating psilocybin-assisted psychotherapy in alcohol use disorder with comorbid depressive symptoms (Luquiens et al., 2025).

Participants were recruited in an inpatient addiction treatment program (CHU Nîmes, France) providing intensive relapse prevention interventions. A total of 30 participants were included following screening of 350 patients. Eligible participants were adults meeting DSM-5 criteria for severe alcohol use disorder and presenting clinically significant depressive symptoms (BDI-II ≥ 14). All participants had completed alcohol detoxification between 14 and 60 days prior to inclusion. Participants were randomly assigned in a 2:1 ratio to receive either a high dose (25 mg; n = 20) or a low dose (1 mg; n = 10) of psilocybin. Randomization was double-blind, with participants, clinicians, and outcome assessors blinded to treatment allocation. The present analyses focus on participants who fully completed the VPT task at both baseline and 3 weeks after the first psilocybin administration (N=25). One participant performed the VPT task at chance level and was therefore excluded from analyses. All participants provided written informed consent prior to inclusion.

### 2.3. Study design and procedure

The study followed the protocol of the PAD trial (Luquiens et al., 2025), which combined psilocybin administration with a structured psychotherapeutic framework including preparatory sessions, a dosing session, and post-session integration. Participants completed the VPT task at two time points: (1) at baseline, prior to psilocybin administration, and (2) 3 weeks after the first psilocybin session. This repeated-measures design allowed the assessment of changes in egocentric and altercentric bias following psilocybin exposure.

The psilocybin administration took place in a controlled clinical setting, with psychological support provided before, during, and after the session in accordance with the study protocol. The post-session VPT assessment was conducted after the acute subjective effects of psilocybin had subsided, in line with the clinical schedule of the PAD trial. Additional clinical and psychological measures (e.g., alcohol use, depressive symptoms, subjective experience) were collected as part of the parent trial. Full details of the therapeutic procedures and assessment timeline are reported elsewhere (Luquiens et al., 2025).

### 2.4. Measures

#### 2.4.1 Visual perspective-taking task

Egocentric bias was assessed using a computerized VPT task adapted from Samson et al. (2010). On each trial, participants were presented with a scene containing an avatar positioned in a room with a variable number of objects visible either from the participant’s perspective or from the avatar’s perspective (see Figure 1). Participants were required to judge either their own perspective (*Self* condition) or the avatar’s perspective (*Other* condition), depending on a cue presented at the beginning of each trial. Trials were further categorized as *congruent* (both perspectives matched) or *incongruent* (the two perspectives differed), allowing the assessment of interference effects. The task consisted of two blocks of 52 trials each, including 48 experimental trials and 4 filler trials per block. Experimental trials were evenly distributed across conditions defined by Perspective (Self vs. Other), Congruency (Congruent vs. Incongruent), and Matching (Match vs. Mismatch). Prior to the experimental blocks, participants completed 26 practice trials with feedback. No reaction time trimming was applied. Trials with incorrect responses were excluded from reaction time analyses. In line with original study, mismatching (expecting “no” answers) and practice trials were discarded from analyses. Egocentric bias was defined as the performance cost associated with judging the avatar’s perspective in incongruent trials relative to congruent trials (i.e., Other/Incongruent minus Other/Congruent), computed separately for reaction times and accuracy. Altercentric bias was defined as the performance cost associated with judging one’s own perspective in incongruent relative to congruent trials (Self/Incongruent minus Self/Congruent). An egocentric superiority index was computed as the difference between egocentric and altercentric bias, where positive values indicate higher egocentric bias than altercentric bias.

**Figure 1.**
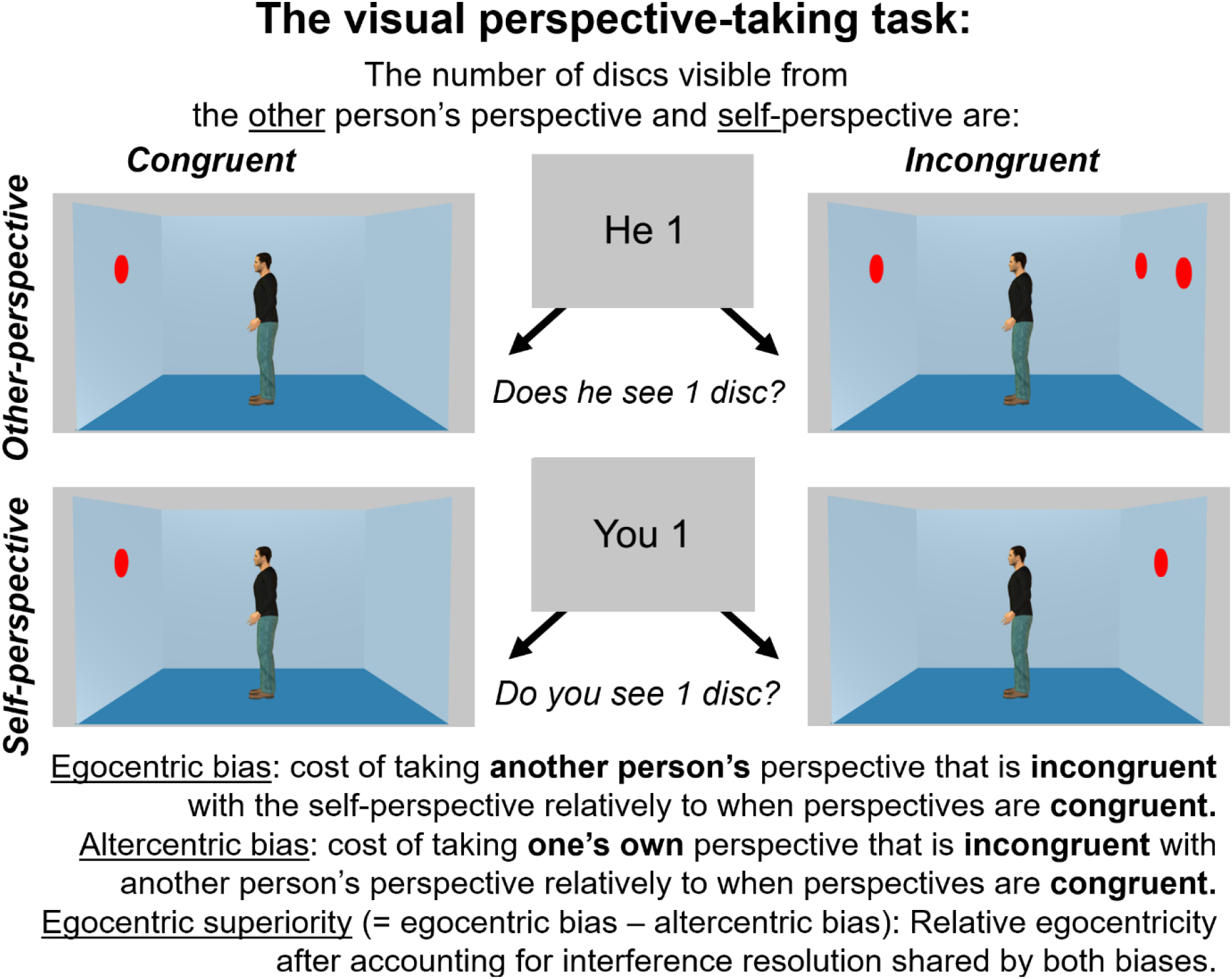
Design and indexes of the level-1 Visual Perspective-Taking task.

#### 2.4.2 Clinical and psychological measures

Demographic variables including sex, age, and education level were collected at baseline as part of the PAD trial protocol.

Subjective psychedelic experiences during the dosing session were assessed using the 5-Dimensions of Altered States of Consciousness questionnaire (5D-ASC; Bodmer et al., 1994). This validated self-report instrument captures key dimensions of the psychedelic experience, including oceanic boundlessness, anxious ego dissolution, and visionary restructuralization, among others. Scores were computed according to standard procedures, with higher scores reflecting more intense subjective experiences (de Deus Pontual et al., 2023). Global cognitive functioning was assessed using the Montreal Cognitive Assessment (MoCA), a widely used screening tool evaluating multiple cognitive domains including attention, executive functions, memory, language, and visuospatial abilities (Alarcon et al., 2015). The MoCA yields a total score ranging from 0 to 30, with higher scores indicating better cognitive functioning.

Post-traumatic stress symptoms were assessed using the PTSD Checklist for DSM-5 (PCL-5), a widely used 20-item self-report measure assessing the severity of post-traumatic stress symptoms experienced during the past month. The questionnaire covers the four DSM-5 PTSD symptom clusters, including intrusion symptoms, avoidance, negative alterations in cognition and mood, and hyperarousal. Items are rated on a 5-point scale ranging from 0 (*Not at all*) to 4 (*Extremely*), with higher scores indicating greater PTSD symptom severity.

Rejection sensitivity was assessed using the Adult Rejection Sensitivity Questionnaire (A-RSQ; Berenson et al., 2009), a self-report measure assessing the tendency to anxiously expect and anticipate rejection in interpersonal situations. Following standard scoring procedures, rejection sensitivity scores were computed across nine hypothetical social scenarios, with higher scores indicating greater rejection sensitivity.

Additional clinical measures (e.g., alcohol use collected with the Alcohol Timeline Followback (Sobell et al., 1996), depressive symptoms assessed with the BDI-II (Beck, 1996)) were collected within the parent trial and are described in detail elsewhere (Luquiens et al., 2025). These measures were not the primary focus of the present analyses.

## 3. Results

### 3.1. Baseline group comparisons

As expected, participants in the high-dose group reported significantly more intense subjective psychedelic experiences than participants in the low-dose group on the 5D-ASC total score, *t*(22) = 2.35, *p* = .028, as well as on the dimensions of Oceanic Boundlessness (*p* = .027), Anxious Ego Dissolution (*p* = .038), Visual Restructuralization (*p* = .011), and Reduction of Vigilance (*p* = .044). Auditory Alterations showed a similar trend (*p* = .053).

The high- and low-dose groups did not differ significantly at baseline on demographic or clinical variables, including sex distribution, χ^2^(1) = 0.23, *p* = .633, age, *t*(22) = -1.60, *p* = .125, cognitive functioning assessed with the MoCA, *t*(22) = 0.85, *p* = .403, post-traumatic stress symptoms assessed with the PCL-5, *t*(22) = 1.15, *p* = .262, or rejection sensitivity assessed with the A-RSQ, *t*(22) = 1.07, *p* = .298.

### 3.2. Effects of psilocybin on perspective-taking performance

We first tested the Time × Dose group interaction separately for each VPT index of performance. For reaction-time egocentric bias, the interaction was significant, *F*(1, 22) = 5.91, *p* = .024, η^2^p = .212. The same was true for the Egocentric Superiority index, *F*(1, 22) = 4.69, *p* = .041, η^2^p = .176.

In the high-dose group (*n* = 18), the egocentric bias and egocentric superiority index significantly decreased from baseline to post-session 1 (*p* = .025, 95% CI [21.94, 164.83]; *p* = .005, 95% CI [76.73, 250.24]). In the low-dose group (*n* = 6), no significant pre-post change was observed (*p* = .246, 95% CI [-229.80, 30.15]; *p* = .547, 95% CI [-195.63, 86.21]).

In short, the significant reductions in egocentric bias and egocentric superiority found in the high-dose group are significantly different from the pre-post changes found the low-dose group (p = .024, 95% CI [-348.86, -27.65], Cohen’s d = -1.15; p = .041, 95% CI [-391.54, - 8.55], Cohen’s d = -1.02).

By contrast, the Time × Dose group interaction was not significant for altercentric bias on reaction times, *F*(1, 22) = 0.03, *p* = .862, η^2^p = .001 and neither on the accuracy-based analyses (egocentric bias: *F*(1, 22) = 1.36, *p* = .256, η^2^p = .058; altercentric bias: *F*(1, 22) = 1.53, *p* = .229, η^2^p = .065; egocentric superiority: *F*(1, 22) = 0.003, *p* = .956, η^2^p < .001).

### 3.3. Exploratory associations with subjective psychedelic experience

Given substantial inter-individual variability in subjective psychedelic experiences, we explored whether 5D-ASC scores were associated with post-session VPT performance. Because the five 5D-ASC dimensions were highly intercorrelated (Cronbach’s α = .978), analyses focused on a composite 5D-ASC total score indexing overall subjective psychedelic intensity. Higher 5D-ASC scores were associated with lower egocentric superiority on accuracy, *r*(23) = −.41, *p* = .049. Associations with egocentric bias reaction times, *r*(23) = −.17, *p* = .426, egocentric superiority on reaction times, *r*(23) = −.19, *p* = .383, and egocentric bias on accuracy, *r*(23) = −.25, *p* = .232, were not significant.

## 4. Discussion

The present study examined via a randomized controlled design whether psilocybin administration modulates egocentric bias during visual perspective taking in individuals with alcohol use disorder (AUD) and persistent depressive symptoms. Participants receiving 25 mg psilocybin showed larger reductions in reaction-time measures of egocentric bias and egocentric superiority than participants receiving 1 mg psilocybin. These effects were specific to egocentric processing, as no effects were observed for altercentric bias. Together, these findings suggest that psilocybin may reduce egocentricity in social cognition three weeks after the dosing session, indicating persisting effects after the acute and afterglow periods.

### 4.1. Psilocybin reduces the dominance of self-perspective in social cognition

The primary finding of this study is that psilocybin selectively reduced egocentric bias in a level-1 visual perspective-taking task. Critically, this effect was observed not only for egocentric bias itself, but also for the egocentric superiority index, which reflects the relative dominance of egocentric over altercentric interference. This pattern indicates that the effect is not explained by a general change in interference resolution or self-other distinction processes that contribute to both egocentric and altercentric biases. If psilocybin had primarily enhanced inhibitory control or self–other distinction mechanisms, comparable effects would be expected on both egocentric and altercentric biases (e.g., Bigot et al., 2025). Instead, it points to a specific reduction in the dominance of the self-perspective.

Consistent with this interpretation, psilocybin did not affect altercentric bias, suggesting that the processing of others’ perspectives was not enhanced. Rather, participants became less constrained by their own perspective, even when controlling for altercentric interference. This distinction is important, as it indicates that persisting effects of psilocybin primarily modulate self-related processing, rather than directly enhancing sensitivity to others. This interpretation can be situated within theoretical accounts proposing that psychedelics transiently reduce the precision or influence of high-level self-representations (Carhart-Harris et al., 2014; Carhart-Harris & Friston, 2019). Within these accounts, the self-perspective can be conceptualized as a high-level prior that typically constrains cognitive processing. In the context of visual perspective taking, egocentric bias reflects the behavioral expression of this constraint, whereby one’s own perspective interferes with the processing of an alternative viewpoint. The reduction in egocentric bias observed following psilocybin administration is therefore consistent with a decrease in the precision of the self-perspective prior, resulting in reduced interference from self-related information. These processes could reflect the persistence of acute effects or differ from processes during the acute experience, described as oceanic boundlessness (Mortaheb et al., 2024) and increased connection with others and emerge later through the psychotherapeutic processed augmented by psilocybin (Wolf et al., 2024).

### 4.2. Egocentric bias as a key mechanism in alcohol use disorder

Egocentric bias provides a useful framework to understand social difficulties in AUD. Prior work has shown that individuals with severe AUD exhibit impairments in tasks traditionally interpreted as theory of mind deficits. However, increasing evidence suggests that these impairments may reflect, at least in part, a combination of self-centred processing and reduced ability to inhibit one’s own perspective, rather than a deficit in mental state inference per se (Maurage et al., 2015; Pabst et al., 2021). Within this framework, the present findings suggest that psilocybin may act on a core socio-cognitive mechanism relevant to AUD, namely the ability to disengage from one’s own perspective in order to process others’ viewpoints. This is particularly relevant given that successful recovery from AUD is strongly linked to the quality of social interactions and relationships, leading to the recently developed concepts of social recovery and social recovery capital, which refers to resources and supports gained through relationships and repeatedly reported to be associated with positive outcomes in addiction (Kaskutas et al., 2002; Kelly et al., 2009, 2012; Francis et al., 2023; Hennessy et al., 2024). A reduction in egocentric bias may therefore facilitate more adaptive interpersonal functioning, potentially contributing to improved clinical outcomes.

### 4.3. Subjective psychedelic experience

The exploratory analyses examining subjective psychedelic experience provide preliminary support for the proposed mechanism. As expected, participants receiving the high dose reported significantly more intense psychedelic experiences than those receiving the low dose, indicating that the 5D-ASC was sensitive to the experimental manipulation. Furthermore, associations between subjective psychedelic intensity and VPT performance were consistently in the predicted direction, with more intense experiences being associated with lower post-dose egocentricity across all indices. Although only the association reached conventional levels of statistical significance for the egocentric superiority on accuracy, this pattern is noteworthy given the limited sample size and consequent lack of statistical power.

Importantly, the five 5D-ASC dimensions were highly intercorrelated in the present sample (Cronbach’s α = .978; intercorrelations *r*s = .86–.98). This suggests that the different dimensions captured largely overlapping variance and did not provide sufficient differentiation to support meaningful dimension-specific analyses. Consequently, the present findings are most consistent with the interpretation that overall subjective psychedelic intensity, rather than any specific experiential component, may be related to reductions in egocentric processing. Future studies with larger samples should examine whether particular aspects of the psychedelic experience, such as oceanic boundlessness or ego dissolution, show more specific relationships with socio-cognitive outcomes.

### 4.4. Limitations

First, although the randomized design reduces the risk of systematic group differences, our analyses are underpowered. The observed effects should be interpreted cautiously. Replication in larger samples will be necessary to determine the robustness of the reported reductions in egocentric bias. Second, the absence of long-term follow-up on VPT performance prevents conclusions about the durability of the observed effects. Additionally, although the use of reaction-time indices allows for sensitive detection of cognitive processes, it remains indirect, and future studies should complement behavioural measures with neural or ecological assessments of social cognition. Third, these results regard a phenotyped population with severe AUD and persistent depressive after detoxification, with high risk of relapse, and could not be generalized to any AUD. Finally, the study did not include a healthy control group. Consequently, it remains unclear whether the observed reduction in egocentric bias reflects a normalization of socio-cognitive processing in AUD or a more general cognitive effect of psilocybin.

## 5. Conclusion

This study provides initial evidence that psilocybin reduces egocentric bias in individuals with alcohol use disorder. This effect was specific to the dominance of the self-perspective, as it was observed for egocentric interference and its superiority over altercentric interference, but not for altercentric bias. These findings suggest that psilocybin primarily reduces the influence of self-related information on cognition, rather than enhancing sensitivity to others. This interpretation is consistent with theoretical models proposing that psychedelics decrease the precision (i.e., the weighting or influence) of high-level self-representations (Carhart-Harris & Friston, 2019). These preliminary findings identify reduced self-perspective dominance as a promising candidate mechanism through which psilocybin may influence social cognition in alcohol use disorder.

## Abbreviations

sAUD: severe alcohol use disorder
VPT: visual perspective taking

## Statements and Declarations

### Ethical considerations

The PAD trial was conducted in accordance with the Declaration of Helsinki and Good Clinical Practice guidelines. The study was approved by [full name and institution of the Comité de Protection des Personnes; ethics approval number] and authorised by the French National Agency for the Safety of Medicines and Health Products (ANSM; authorisation number to be confirmed). The trial was registered at ClinicalTrials.gov (NCT06235411).

### Consent to participate

All participants provided written informed consent before enrolment in the PAD trial.

### Consent for publication

Not applicable. The manuscript does not contain identifiable information or images of individual participants.

## Data availability

The participant-level data are not publicly available because they contain sensitive clinical information. Requests for access may be directed to the corresponding author and will be considered subject to the applicable ethical approvals and data protection requirements.

